# Comparative Analysis of Long COVID and Post-Vaccination Syndrome: A Cross-Sectional Study of Clinical Symptoms and Machine Learning-Based Differentiation

**DOI:** 10.1101/2025.08.14.25333639

**Authors:** Harlan M. Krumholz, Mitsuaki Sawano, Yilun Wu, Rishi Shah, Tianna Zhou, Adith S. Arun, Pavan Khosla, Shayaan Kaleem, Anushree Vashist, Bornali Bhattacharjee, Yuan Lu, Frederick Warner, Chenxi Huang, Leying Guan, César Caraballo, David Putrino, Danice Hertz, Brianne Dressen, Teresa Michelsen, Liza Fisher, Cynthia Adinig, Pamela Bishop, Akiko Iwasaki

## Abstract

**Importance:** Long COVID is a well-documented post-viral syndrome, while post-vaccination syndrome (PVS) remains poorly characterized. Understanding their similarities and differences is essential for refining diagnostic criteria and developing targeted interventions. This study systematically compares the symptomatology of long COVID and PVS following COVID-19 vaccination, highlighting key distinctions that could inform clinical practice and research.

**Objective:** To assess the clinical characteristics of long COVID and PVS and identify key distinguishing features between the conditions.

**Design, Setting and Participants:** This cross-sectional analysis used questionnaire data from the decentralized Yale Listen to Immune, Symptom and Treatment Experiences Now (LISTEN) Study, collected from May 2022 to July 2023. Data analysis occurred between July 2023 and May 2024. A convenience sample of adults (age ≥18 years) with either long COVID or PVS was included.

**Main Outcomes and Measures:** Symptom data were analyzed using clustering techniques to identify groups with shared symptom patterns. A gradient-boosted machine learning model was used to determine the most distinguishing symptoms between long COVID and PVS.

**Results:** The long COVID group (n = 441) and PVS group (n = 241) had similar demographic profiles (median age 46 years; 74% vs 80% female, respectively). Participants with long COVID most commonly reported brain fog, altered sense of smell and taste, shortness of breath, fatigue, memory problems, and difficulty speaking. Participants with PVS more frequently reported burning sensations, neuropathy, and numbness. Clustering analysis identified three symptom-based subgroups: one enriched for neurological symptoms and PVS; one characterized by multi-system symptoms and predominantly long COVID; and one dominated by psychiatric and sleep symptoms, also primarily long COVID. The machine learning model achieved an AUC of 0.79 (95% CI, 0.75–0.82) and highlighted altered sense of smell, cough, burning sensations, and brain fog as key differentiators.

**Conclusions and Relevance:** Although long COVID and PVS share overlapping symptoms, they have distinct clinical profiles, suggesting the possibility of different underlying biological mechanisms. These distinctions may help refine diagnostic criteria, guide personalized treatment strategies, and inform further research into their respective pathophysiology.

**KEY POINTS:** *Question:* What are the similarities and differences between long COVID and post-vaccination syndrome (PVS)?

*Findings:* In this cross-sectional study of 682 individuals, machine learning models identified distinct symptoms between long COVID and PVS. Long COVID was characterized by brain fog, altered sense of smell, and shortness of breath, while PVS was associated with burning sensations, neuropathy, and numbness.

*Meaning:* Although long COVID and PVS share overlapping symptoms, they have distinctive symptom profiles, suggesting potentially different underlying biological mechanisms. Understanding these differences can guide clinical diagnosis and targeted management, and inform further research into their distinct immune and biological pathways.

## INTRODUCTION

Post-vaccination syndrome (PVS) refers to chronic, debilitating symptoms that emerge shortly after receiving a vaccine—a phenomenon that has received increasing attention following widespread COVID-19 vaccination.^1-11^ Unlike the typical short-term side effects that resolve within days, PVS involves persistent symptoms that may significantly impair quality of life. Long COVID, or post-acute sequelae of SARS-CoV-2 infection (PASC), is another post-exposure condition characterized by prolonged, multisystem symptoms following SARS-CoV-2 infection.

Both PVS and long COVID share features of chronic illness and broad symptomatology. Importantly, both may result from exposure to the SARS-CoV-2 spike protein—via infection or vaccination—but whether they share a common underlying pathophysiology remains uncertain. Some researchers have proposed that persistent immune dysregulation or inflammation triggered by the spike protein could contribute to both conditions.^12^

Comparing the symptom profiles of long COVID and PVS may provide insight into shared and divergent mechanisms, improving our understanding of both conditions. Accordingly, we analyzed symptom patterns and demographic characteristics in individuals self-identifying as having either long COVID or PVS. We aimed to assess the onset, severity, and distribution of symptoms and to evaluate whether symptom profiles can distinguish the two syndromes.

## METHODS

### Study Design

The Yale Listen to Immune, Symptom and Treatment Experiences Now (LISTEN) study is a decentralized, participant-centric observational study of adults experiencing long COVID and PVS. The LISTEN study, launched in May 2022, recruited participants via Hugo Health Kindred, an online patient community of individuals interested in contributing to research on these conditions. Participants consented to share survey data regarding symptoms and diagnoses. This analysis specifically focuses on symptom and diagnostic data obtained via structured surveys.

### Study Population

Participants were adults (age ≥18 years) who self-reported experiencing either long COVID or PVS. To clearly differentiate syndromes, participants who reported having both long COVID and PVS were excluded from the primary analysis (**eFigure 1**). Long COVID and PVS were defined strictly by participants’ self-report, as there are no validated biomarkers.

### Data Collection

Participants completed online surveys through Hugo Health Kindred, reporting demographics, vaccination history, acute COVID-19 infection history, comorbid conditions, and symptoms. Surveys were iteratively refined based on patient input to enhance clarity, reduce participant burden, and ensure relevance. Patient research partners (DH, LF) contributed to the survey’s development, and additional patient partners (LF, CA, BD, PB) participated in interpreting the findings.

Symptoms were captured using an extensive checklist (96 symptoms), with questions explicitly linked to either long COVID or PVS onset. Participants also rated their current health status using the Euro-QoL visual analogue scale (EQ-VAS; 0-100) and symptom severity using a 0-100 scale indicating symptom severity on their worst days. Self-reported health status was assessed on a 5-point scale (excellent, very good, good, fair, or poor) based on an individual’s self-perceived general health. We also included the question: “On your worst days, how bad are your symptoms (0 being a trivial illness and 100 being unbearable)?”

### Statistical Analysis

We described participant characteristics using median and interquartile ranges for continuous variables and proportions for categorical variables. Comparisons between long COVID and PVS groups used Chi-squared or Fisher’s exact tests for categorical variables, and Wilcoxon rank-sum tests for continuous variables. We used Bonferroni correction to adjust for multiple comparisons.

We used co-clustering analysis employing a bipartite spectral clustering method to identify symptom clusters shared across both long COVID and PVS groups.^13^ To simplify the analysis, symptoms were first grouped into 10 clinically meaningful categories. K-means clustering was applied, and the optimal number of clusters was determined using the Calinski-Harabasz Index.^14^ Cluster stability was evaluated through subsampling (90% subsamples, repeated 1000 times).

To differentiate long COVID from PVS, a gradient-boosted tree machine learning model was developed, trained using symptom presence or absence as predictors.^15, 16^ Model performance was summarized by the area under the receiver operating characteristic curve (AUROC). The most discriminative symptoms were identified using permutation-based variable importance metrics,^17^ progressively excluding less important variables until a significant performance drop (>1.5% AUROC decrease) occurred. Robustness was confirmed using alternative machine learning algorithms (XGBoost Gain, XGBoost SHAP values).^18, 19^

### Ethical Considerations

The Yale University Institutional Review Board approved the LISTEN study, which conforms to the Declaration of Helsinki and STrengthening the Reporting of OBservational studies in Epidemiology (STROBE) reporting guidelines. Dr. Krumholz, a co-founder of Hugo Health, developed the Hugo Kindred platform. His involvement in this study was overseen by the Yale Conflict of Interest Committee.

## RESULTS

### Study Sample

The study sample consisted of 441 participants with long COVID and 241 with PVS (**Table 1**), with surveys completed between May 2022 and July 2023. Participants with long COVID completed surveys at a median of 447 days (IQR: 225–784 days) post-index infection, while PVS participants did so at a median of 595 days (IQR: 417–661 days; range: 40–1058 days) post-vaccination. Both groups had similar median ages (46 years) and were predominantly female (74% long COVID, 80% PVS). There were no significant differences in racial or ethnic composition, marital status, employment, income, health insurance, or social support measures (**Table 1**). However, long COVID participants were significantly more likely to reside in the United States (p=0.03).

**Table 1.**
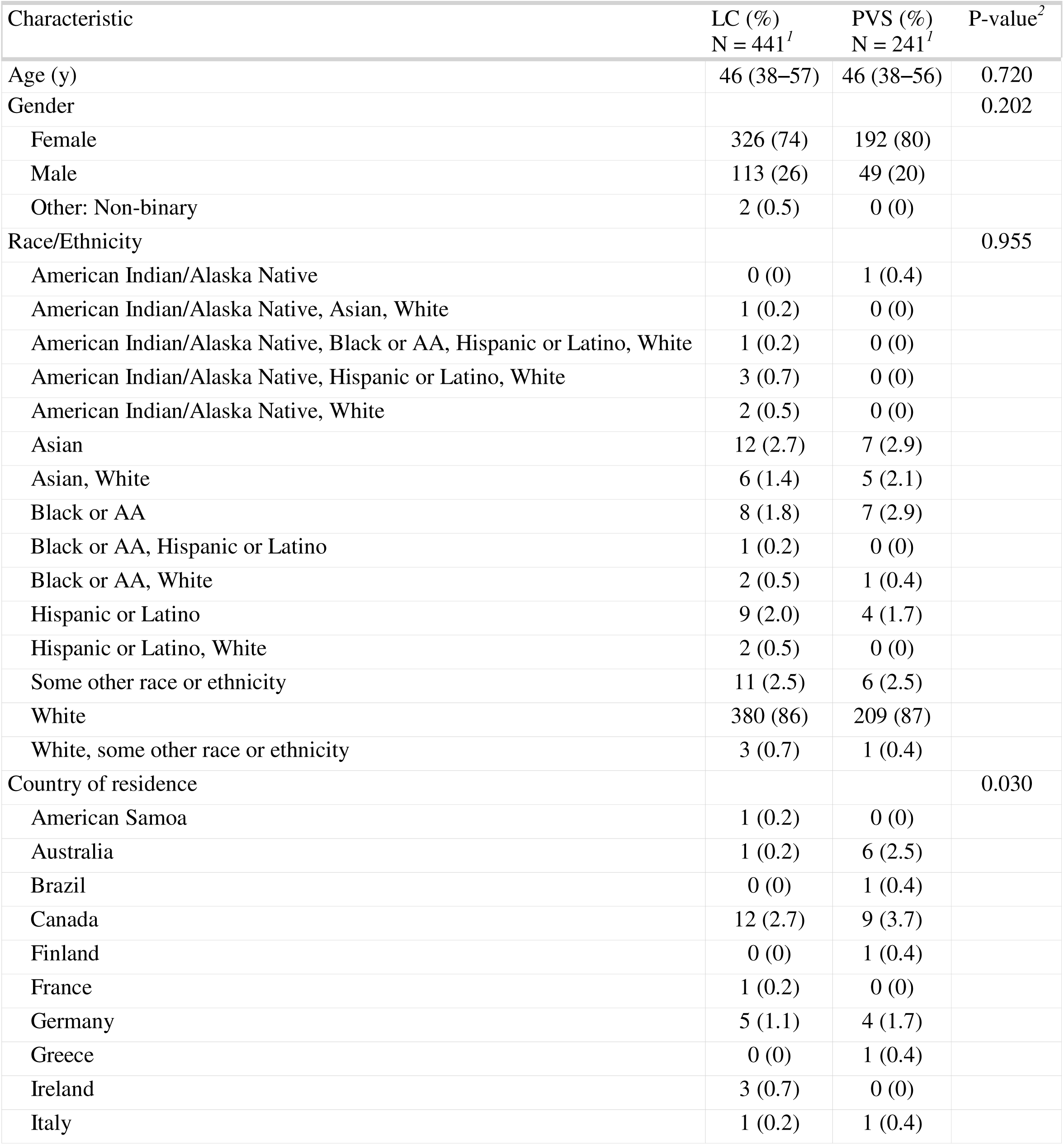
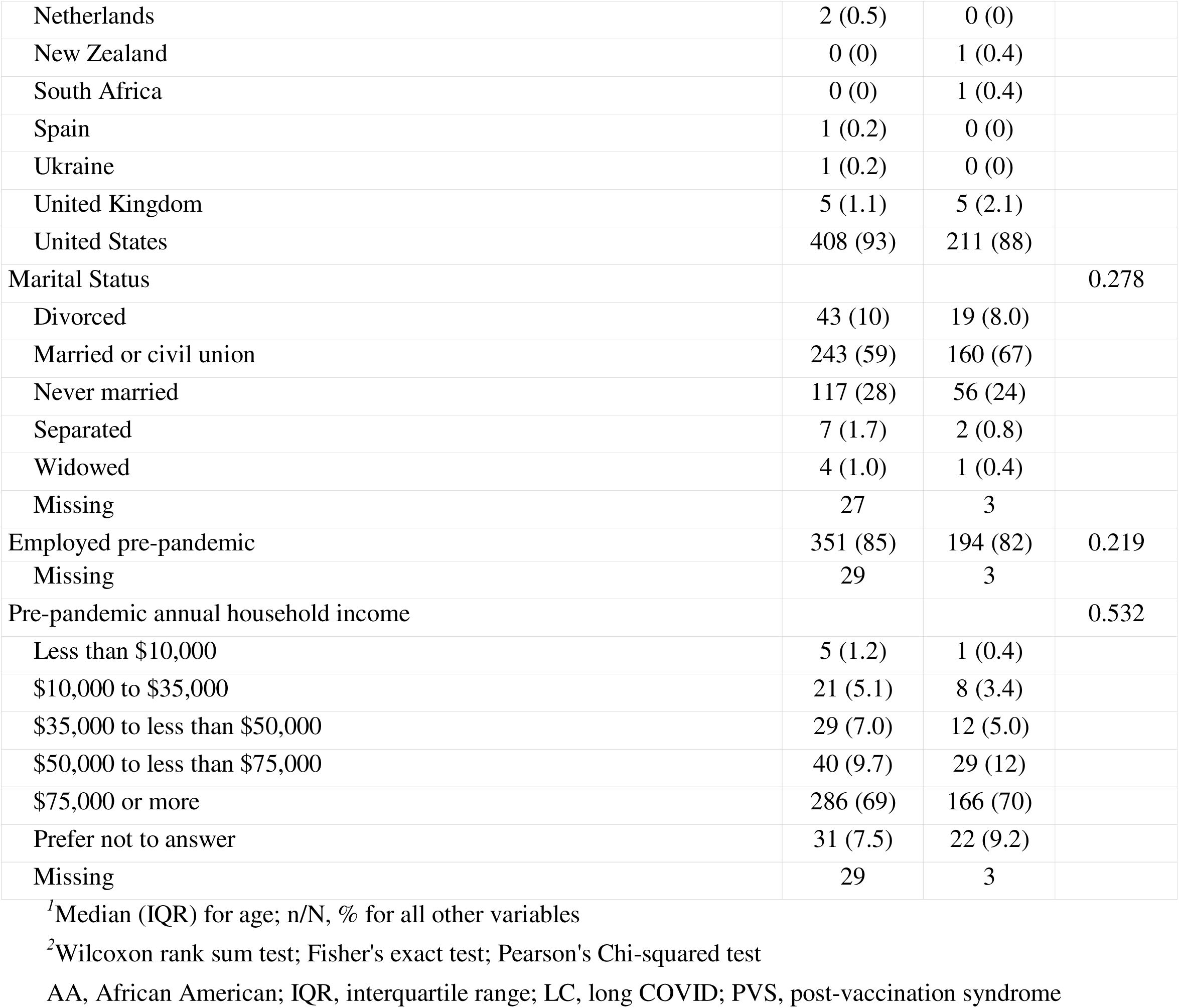
Participant demographic and socioeconomic characteristics.

| Characteristic | LC (%)<br>N = 441<sup>1</sup> | PVS (%)<br>N = 241<sup>1</sup> | P-value<sup>2</sup> |
| --- | --- | --- | --- |
| Age (y) | 46 (38–57) | 46 (38–56) | 0.720 |
| Gender |  |  | 0.202 |
| Female | 326 (74) | 192 (80) |  |
| Male | 113 (26) | 49 (20) |  |
| Other: Non-binary | 2 (0.5) | 0 (0) |  |
| Race/Ethnicity |  |  | 0.955 |
| American Indian/Alaska Native | 0 (0) | 1 (0.4) |  |
| American Indian/Alaska Native, Asian, White | 1 (0.2) | 0 (0) |  |
| American Indian/Alaska Native, Black or AA, Hispanic or Latino, White | 1 (0.2) | 0 (0) |  |
| American Indian/Alaska Native, Hispanic or Latino, White | 3 (0.7) | 0 (0) |  |
| American Indian/Alaska Native, White | 2 (0.5) | 0 (0) |  |
| Asian | 12 (2.7) | 7 (2.9) |  |
| Asian, White | 6 (1.4) | 5 (2.1) |  |
| Black or AA | 8 (1.8) | 7 (2.9) |  |
| Black or AA, Hispanic or Latino | 1 (0.2) | 0 (0) |  |
| Black or AA, White | 2 (0.5) | 1 (0.4) |  |
| Hispanic or Latino | 9 (2.0) | 4 (1.7) |  |
| Hispanic or Latino, White | 2 (0.5) | 0 (0) |  |
| Some other race or ethnicity | 11 (2.5) | 6 (2.5) |  |
| White | 380 (86) | 209 (87) |  |
| White, some other race or ethnicity | 3 (0.7) | 1 (0.4) |  |
| Country of residence |  |  | 0.030 |
| American Samoa | 1 (0.2) | 0 (0) |  |
| Australia | 1 (0.2) | 6 (2.5) |  |
| Brazil | 0 (0) | 1 (0.4) |  |
| Canada | 12 (2.7) | 9 (3.7) |  |
| Finland | 0 (0) | 1 (0.4) |  |
| France | 1 (0.2) | 0 (0) |  |
| Germany | 5 (1.1) | 4 (1.7) |  |
| Greece | 0 (0) | 1 (0.4) |  |
| Ireland | 3 (0.7) | 0 (0) |  |
| Italy | 1 (0.2) | 1 (0.4) |  |
| Netherlands | 2 (0.5) | 0 (0) |  |
| New Zealand | 0 (0) | 1 (0.4) |  |
| South Africa | 0 (0) | 1 (0.4) |  |
| Spain | 1 (0.2) | 0 (0) |  |
| Ukraine | 1 (0.2) | 0 (0) |  |
| United Kingdom | 5 (1.1) | 5 (2.1) |  |
| United States | 408 (93) | 211 (88) |  |
| Marital Status |  |  | 0.278 |
| Divorced | 43 (10) | 19 (8.0) |  |
| Married or civil union | 243 (59) | 160 (67) |  |
| Never married | 117 (28) | 56 (24) |  |
| Separated | 7 (1.7) | 2 (0.8) |  |
| Widowed | 4 (1.0) | 1 (0.4) |  |
| Missing | 27 | 3 |  |
| Employed pre-pandemic | 351 (85) | 194 (82) | 0.219 |
| Missing | 29 | 3 |  |
| Pre-pandemic annual household income |  |  | 0.532 |
| Less than \$10,000 | 5 (1.2) | 1 (0.4) | |
| \$10,000 to \$35,000 | 21 (5.1) | 8 (3.4) | |
| \$35,000 to less than \$50,000 | 29 (7.0) | 12 (5.0) | |
| \$50,000 to less than \$75,000 | 40 (9.7) | 29 (12) | |
| \$75,000 or more | 286 (69) | 166 (70) | |
| Prefer not to answer | 31 (7.5) | 22 (9.2) |  |
| Missing | 29 | 3 |  |

<sup>1</sup>Median (IQR) for age; n/N, % for all other variables
<sup>2</sup>Wilcoxon rank sum test; Fisher's exact test; Pearson's Chi-squared test
AA, African American; IQR, interquartile range; LC, long COVID; PVS, post-vaccination syndrome

Eighty-eight percent of long COVID participants reported receiving at least one COVID-19 vaccination (**Table 2**). Roughly half of the participants in both groups reported having pre-existing medical conditions affecting their health (52% long COVID, 50% PVS) (**eTable 1**). Self-reported health status of excellent or very good was similar between groups (24% long COVID, 27% PVS). Median EQ-VAS scores were comparable at 49 (IQR: 32–61) for long COVID and 50 (IQR: 39–70) for PVS (**eFigure 2**).

**Table 2.**
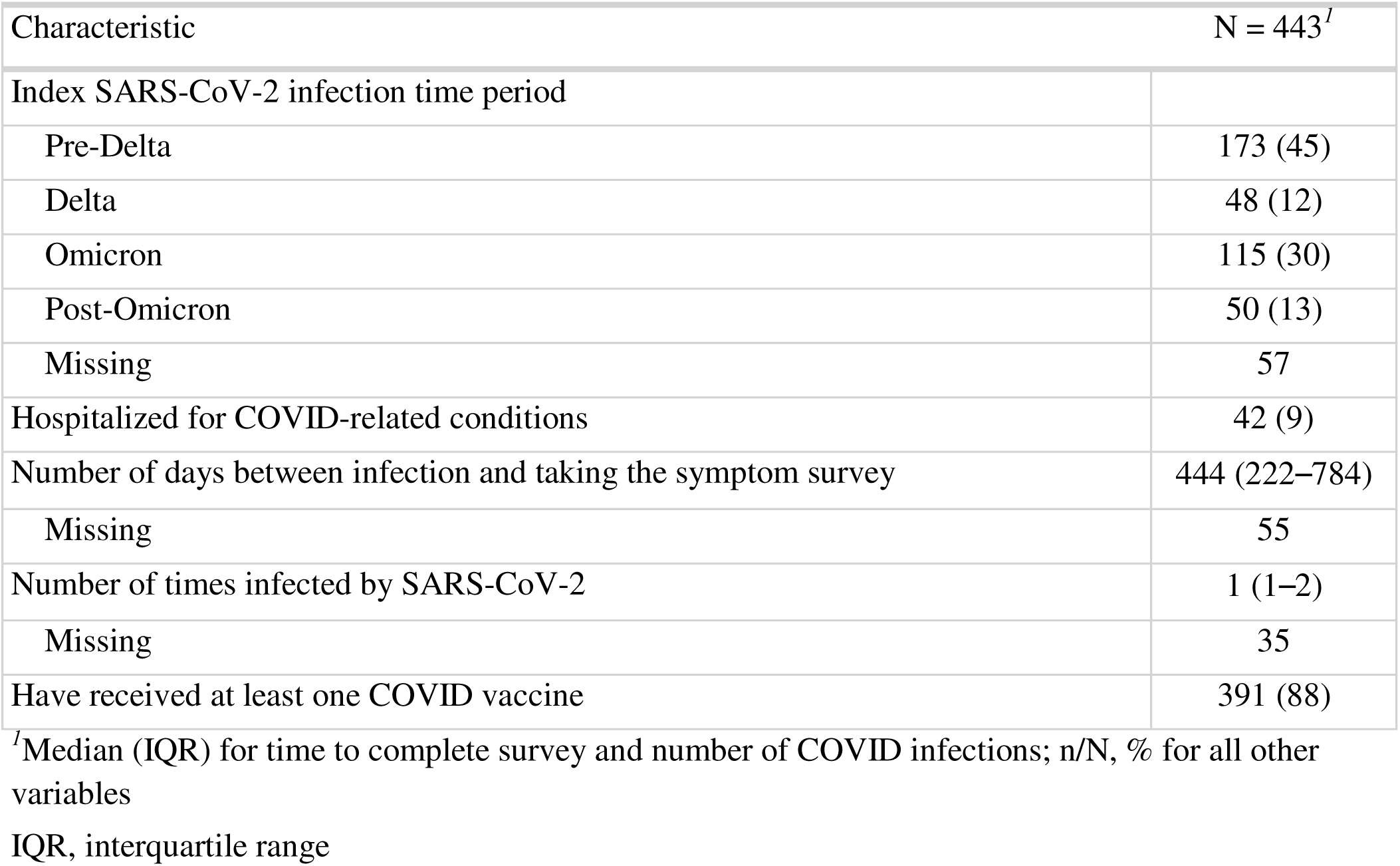
SARS-CoV-2 infection characteristics among participants with long COVID.

### Timing of the Index Acute COVID Infection/Vaccination and Symptom Onset

The majority (45%) of long COVID participants reported their index infection during the Pre-Delta COVID-19 wave (**Table 2**). PVS participants most frequently reported receiving the Pfizer-BioNTech vaccine (55%) followed by Moderna (37%). Symptoms began after the first vaccine dose in 44%, after the second dose in 33%, and after third or subsequent doses in 23%. Symptom onset occurred a median of 3 days post-vaccination (IQR: 1–8 days; **Table 3**).

**Table 3.**
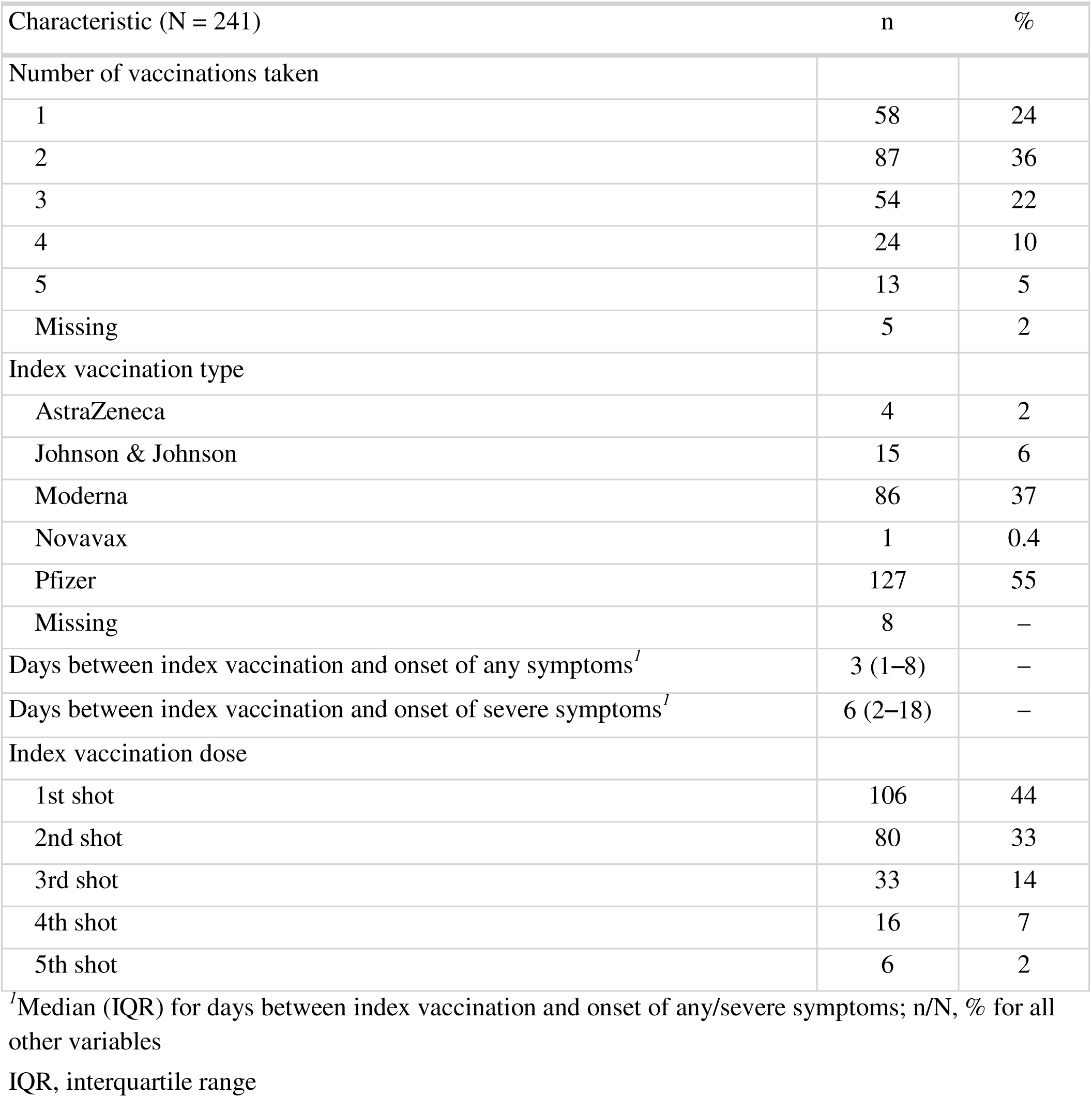
Vaccinations and timing of symptom onset of PVS.

| Characteristic (N = 241) | n | % |
| --- | --- | --- |
| Number of vaccinations taken |  |  |
| 1 | 58 | 24 |
| 2 | 87 | 36 |
| 3 | 54 | 22 |
| 4 | 24 | 10 |
| 5 | 13 | 5 |
| Missing | 5 | 2 |
| Index vaccination type |  |  |
| AstraZeneca | 4 | 2 |
| Johnson & Johnson | 15 | 6 |
| Moderna | 86 | 37 |
| Novavax | 1 | 0.4 |
| Pfizer | 127 | 55 |
| Missing | 8 | – |
| Days between index vaccination and onset of any symptoms<sup>1</sup> | 3 (1–8) | – |
| Days between index vaccination and onset of severe symptoms<sup>1</sup> | 6 (2–18) | – |
| Index vaccination dose |  |  |
| 1st shot | 106 | 44 |
| 2nd shot | 80 | 33 |
| 3rd shot | 33 | 14 |
| 4th shot | 16 | 7 |
| 5th shot | 6 | 2 |

<sup>1</sup>Median (IQR) for days between index vaccination and onset of any/severe symptoms; n/N, % for all other variables
IQR, interquartile range

### Health Status and Symptom Frequencies

Among the people with long COVID, 54% described their health as fair or poor; and among those with PVS, 44% described their health as fair or poor. As measured by the EQ-VAS, the median score was 49 (IQR: 32–61) in the long COVID group compared with 50 (IQR: 39–70) in the PVS group. Symptom severity on worst days was similar between groups (median score of 79 for long COVID vs. 80 for PVS; **eTable 2, eFigure 2).**

Participants’ symptoms are shown in **Table 4** and **eFigure 3**. Among long COVID participants, excessive fatigue was the most common symptom (87%), whereas exercise intolerance was most prevalent in PVS participants (71%). Participants with long COVID reported significantly higher prevalence of brain fog (p<0.0001), shortness of breath (p<0.0001), excessive fatigue (p<0.0001), altered sense of smell (p<0.0001), altered sense of taste (p<0.0001), and memory problems (p<0.0001). Participants with PVS were significantly more likely to report burning sensations (p<0.0001) and neuropathy (p=0.001).

**Table 4.**
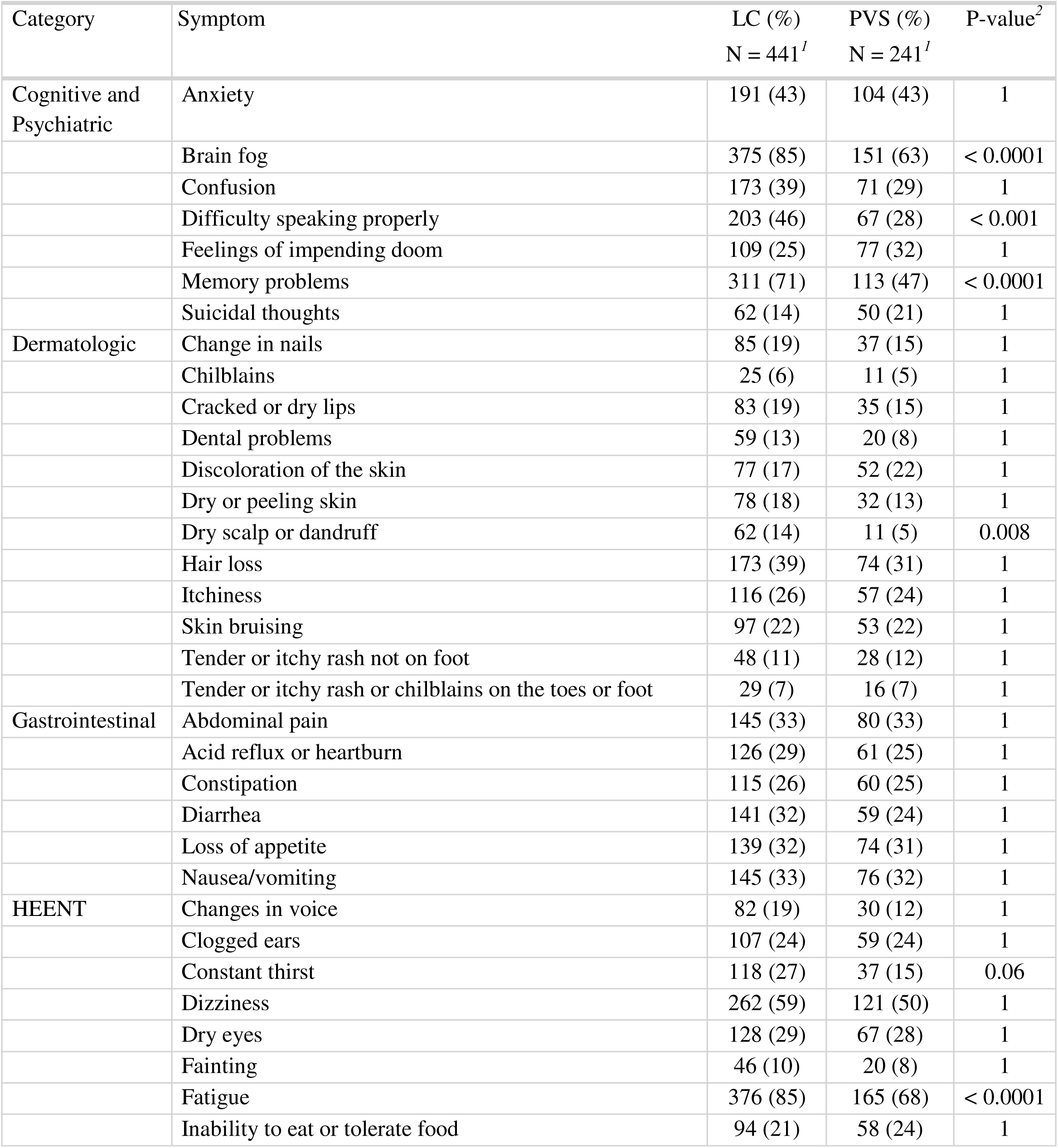
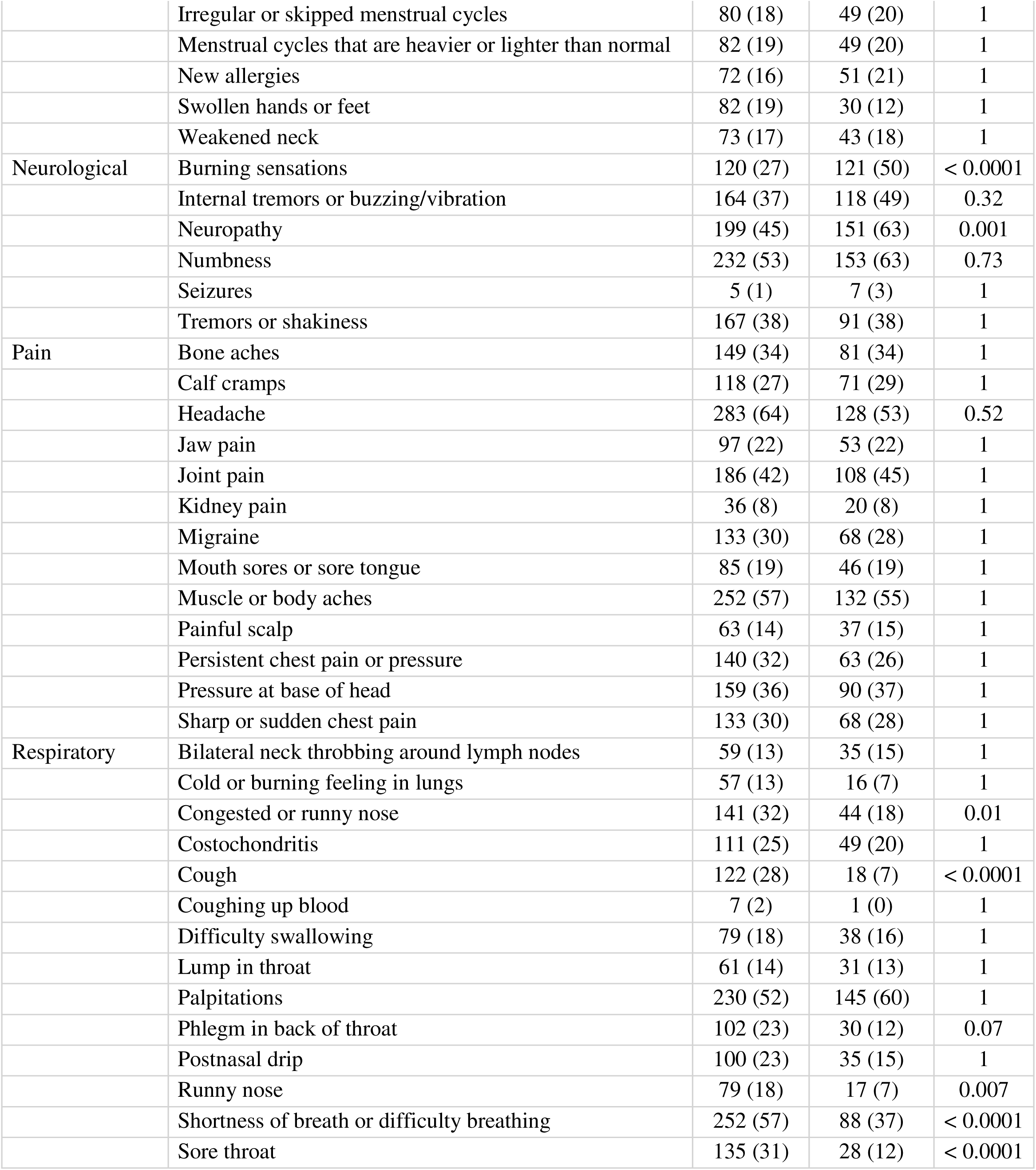
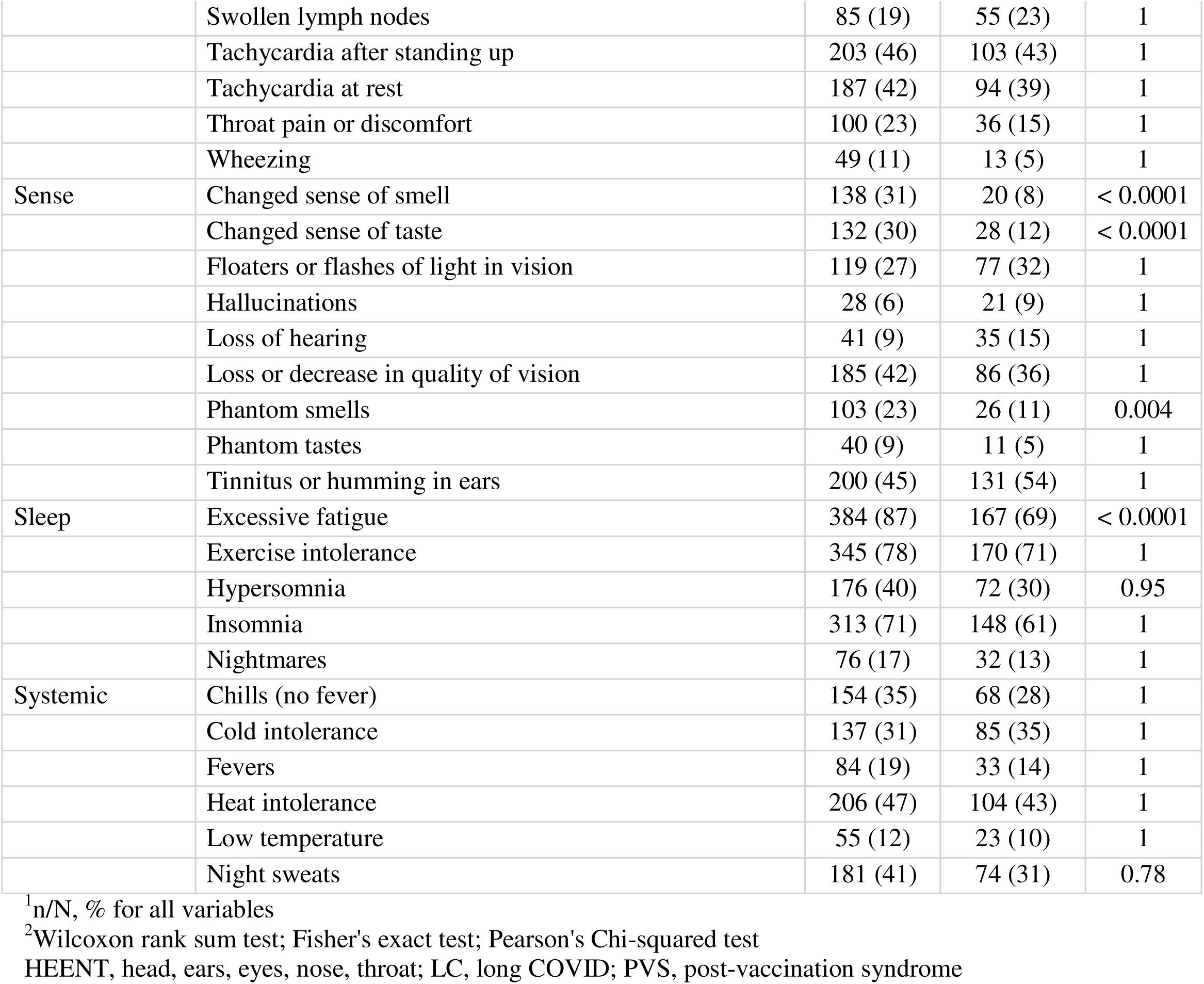
Participant symptoms.

### New Diagnoses and Treatment

Since the pandemic began, participants with PVS reported significantly more new diagnoses of neurologic conditions compared with participants with long COVID (p<0.0001; **eTable 3**). Participants with long COVID more frequently reported using antidiarrheals (p=0.044), bronchodilators (p<0.0001), inhaled steroids (p<0.0001), and nirmatrelvir (p=0.001), and having received speech-related physical therapy (p=0.013) compared with those with PVS. In contrast, participants with PVS more often reported using black seed oil (p=0.003), dandelion (p=0.005), intermittent fasting (p=0.016), eastern white pine needle (p<0.001), and other oral medications (p<0.001) than those with long COVID (**eTable 4, eFigure 4**).

### Clustering Analysis

Symptom clustering analysis identified three distinct symptom clusters (**Figure 1**) among participants. Cluster A (7.4% of all participants, 72% PVS) primarily included neurological symptoms. Cluster B (58.0% of all participants, 65.7% long COVID) exhibited widespread symptoms across the categories of Pain; Head, Eyes, Ears, Nose, and Throat (HEENT); Respiratory; Gastrointestinal; Systemic; Dermatologic; and Sense. Cluster C (34.6% of all participants, 71.5% long COVID) featured predominantly psychiatric and sleep-related symptoms. Cluster stability was confirmed through repeated subsampling, demonstrating an average overlap of 90.7% to 97.6% across clusters.

**Figure 1.** Clustering of participants with prevalence of symptom categories. Participants in cluster A were mainly from the PVS group and had mostly neurological symptoms. Participants in cluster B were predominantly from the long COVID group and had widespread symptoms among all categories. Participants in cluster C were primarily from the long COVID group and had a majority of psychiatric and sleep-related symptoms. HEENT, Head, Eyes, Ears, Nose, and Throat; PVS, post-vaccination syndrome

### Differentiating Individuals with Long COVID and PVS

A gradient-boosted machine learning model successfully distinguished participants with long COVID from those with PVS (AUC = 0.79; 95% CI: 0.75–0.82) **(Figure 2)**. The most important discriminative symptoms were altered sense of smell, cough, burning sensation, and brain fog. Consistency of these findings was validated using alternative machine learning methods (XGBoost Gain, XGBoost SHAP), which showed high concordance (Pearson correlations 0.95–0.98, p<0.001).

**Figure 2.** A gradient-boosted machine learning model identifies seven symptoms that help differentiate patients experiencing long COVID from those experiencing post-vaccination syndrome (PVS). A) Receiver operating curve for the gradient-boosted machine learning model with seven symptoms in distinguishing patients experiencing long COVID from those experiencing PVS. Area under the curve is 0.79 with 95% confidence interval of 0.75 to 0.82. B) Variable importance values for each of the seven symptoms in the gradient-boosted machine learning model and the percent importance of each variable in differentiating participants with long COVID as opposed to PVS. AUC, area under the curve; PVS, post-vaccination syndrome

The probability distribution based on the model highlighted symptom enrichment differences between long COVID and PVS. Participants in the highest decile of long COVID probability predominantly experienced cough and altered senses of smell and taste, while those in the lowest decile (indicative of PVS) commonly reported tingling, neuropathy, and burning sensations (**Figure 3**).

**Figure 3.** Symptom prevalence for sufficiently different symptoms (p < 0.05 with Wilcoxon two-sample test) between the first and tenth decile of predicted probability of having long COVID based on the gradient-boosted machine learning model.

## DISCUSSION

This study indicates that long COVID and PVS, while sharing common features, are distinguishable conditions based on their clinical symptom profiles. Using clustering and machine learning approaches, we identified specific symptom phenotypes—such as brain fog, altered smell, cough, and burning sensations—that most effectively differentiated the two groups. These findings suggest potentially distinct underlying biological mechanisms, which could help refine diagnostic criteria and guide targeted management strategies.

In bivariate analyses, participants with PVS were significantly more likely to report neuropathic symptoms, including burning sensations and numbness. In contrast, long COVID was more commonly associated with respiratory and sensory symptoms, such as brain fog, shortness of breath, and altered senses of smell and taste. These findings expand current understanding by providing a more granular description of symptom differences—information that is critical for clinical awareness and patient care.

Clustering analysis revealed three distinct symptom-based subgroups, including one enriched for PVS and dominated by neurological symptoms. This supports the hypothesis that while both conditions may involve immune responses to the SARS-CoV-2 spike protein, additional mechanisms—such as direct viral tissue effects—may be more relevant in long COVID. For example, disturbances in smell and taste may reflect local infection of olfactory and gustatory tissues.^20-22^ The ability to distinguish these syndromes based on symptomatology reinforces the notion that they may arise, at least in part, through different biological pathways.^23^

Although these findings are promising, the symptom-based classification remains exploratory and requires further validation. Some individuals with long COVID may share pathophysiological features with PVS, while others may not. Future research should prioritize longitudinal designs and integrate immune phenotyping to assess whether distinct symptom clusters correlate with underlying biological signatures.

Our results also underscore the clinical complexity of differentiating these syndromes in practice. Patients may present with overlapping features, and some individuals with PVS may have had unrecognized SARS-CoV-2 infection—especially early in the pandemic when testing was limited. Conversely, patients with long COVID may under-report vaccine-related symptoms due to limited public discourse around non-acute vaccine effects. These diagnostic ambiguities highlight the need for validated biomarkers and greater clinical awareness of both conditions. Also, future research should prioritize longitudinal studies and immune phenotyping to validate findings, clarify mechanisms, and improve management strategies. The clear symptom differentiation identified provides an important step towards personalized treatment for individuals with long COVID and PVS.

This study has several limitations. It relies on self-reported diagnoses and symptoms, without verification through medical records. Participation was voluntary and online, potentially introducing self-selection bias. Additionally, the study was conducted in English, which may have limited participant diversity. As a descriptive analysis, it was not designed to determine the incidence or etiology of either condition.

In conclusion, long COVID and PVS appear to be distinct syndromes with partially overlapping but differentiable symptom profiles. Both deserve clinical and scientific attention, given their burden on affected individuals. Our findings support the need for integrated clinical and biological research to unravel the mechanisms underlying these conditions and to develop tailored diagnostic and therapeutic approaches. Symptom differentiation may be a foundation for future precision medicine strategies targeting long COVID and PVS.

## Data Availability

All data produced in the present study are available upon reasonable request to the corresponding author.

## Funding Statement

This study was funded in part by the Howard Hughes Medical Institute Collaborative COVID-19 Initiative, and in part by Fred Cohen and Carolyn Klebanoff.

## Conflict of Interest Disclosures

H.M.K., in the past three years, received options for Element Science and Identifeye and payments from F-Prime for advisory roles. He was a co-founder of and held equity in Hugo Health. He is a co-founder of and holds equity in Refactor Health and ENSIGHT-AI. He is associated with research contracts through Yale University from Janssen, Kenvue, Novartis, and Pfizer. B.B. was partially supported by, and C.C. was supported by, a grant from the Yale-Mayo Clinic Center of Excellence in Regulatory Science and Innovation (CERSI) (U01FD005938). A.I. co-founded RIGImmune, Xanadu Bio and PanV and is a member of the Board of Directors of Roche Holding and Genentech. B.D. reports being a plaintiff in a lawsuit against AstraZeneca alleging breach of contract following her volunteer participation in 2020 in their COVID-19 vaccine clinical trial. She is also a co-chair of REACT19, a non-profit organization offering financial, physical, and emotional support for those suffering from long-term COVID-19 vaccine adverse events. D.H. serves on the Advisory Board of REACT19.

## Data Sharing Statement

The data supporting this study’s findings are available from the corresponding author, H.M.K., upon reasonable request and funding for deidentification.

## Notes

### Author Declarations

The Yale University Institutional Review Board approved the LISTEN study.

