## Supplemental Materials for "Comparative Analysis of Long COVID and Post-Vaccination Syndrome: A Cross-Sectional Study of Clinical Symptoms and Machine Learning-Based Differentiation"

**SUPPLEMENTAL MATERIAL**

**eTables**

eTable 1. Pre-pandemic comorbid conditions.

eTable 2. Participants’ health status.

eTable 3. New-onset conditions.

eTable 4. New treatments.

**eFigures**

eFigure 1. Study flow diagram.

eFigure 2. Distribution of health status.

eFigure 3. Differences in reported symptoms between long COVID and PVS.

eFigure 4. Differences in treatment use between long COVID and PVS.

**eTable 1. Pre-pandemic comorbid conditions.**

| **Pre-existing Condition** | **LC (%)**  **N = 441*^1^*** | **PVS (%)**  **N = 241*^1^*** | **P-value*^2^*** |
| --- | --- | --- | --- |
| Any allergies | 215 (49) | 116 (48) | 1 |
| Arthritis (including rheumatoid arthritis, gout, lupus, or fibromyalgia) | 74 (17) | 31 (13) | 1 |
| Asthma | 84 (19) | 49 (20) | 1 |
| Autoimmune disease (including lupus, scleroderma, etc.) | 51 (12) | 26 (11) | 1 |
| Bleeding disorder (including sickle cell disease or Thalassemia) | 4 (1) | 3 (1) | 1 |
| Blood clots | 7 (2) | 2 (1) | 1 |
| Cancer or malignancy of any kind | 22 (5) | 13 (5) | 1 |
| Cerebrovascular conditions affecting blood vessels to or in the brain (including stroke) | 4 (1) | 2 (1) | 1 |
| Chronic lung disease (including emphysema, chronic bronchitis, chronic obstructive pulmonary disease (COPD), or pulmonary fibrosis) | 11 (2) | 3 (1) | 1 |
| Cystic fibrosis | 0 (0) | 0 (0) | 1 |
| Diabetes | 11 (2) | 6 (3) | 1 |
| Ehlers-Danlos Syndrome (hypermobile joints) | 10 (2) | 8 (3) | 1 |
| Gastrointestinal issues (including IBS or acid reflux) | 109 (25) | 67 (28) | 1 |
| Heart attack, also called myocardial infarction | 3 (1) | 2 (1) | 1 |
| Heart conditions (including coronary artery disease or cardiomyopathies) | 13 (3) | 8 (3) | 1 |
| Heart failure | 1 (0) | 0 (0) | 1 |
| High cholesterol | 65 (15) | 34 (14) | 1 |
| History of organ transplant (including kidney, liver, heart, or lung) | 0 (0) | 0 (0) | 1 |
| Hypertension or high blood pressure | 53 (12) | 31 (13) | 1 |
| Immunocompromised state (including weakened immune system from blood or bone marrow transplant, immune deficiencies, HIV, use of corticosteroids, or use of other immune-weakening medicines) | 14 (3) | 6 (3) | 1 |
| Kidney disease | 5 (1) | 1 (0) | 1 |
| Liver disease | 3 (1) | 0 (0) | 1 |
| Lyme disease | 27 (6) | 7 (3) | 1 |
| MCAS (mast cell activation syndrome) or other mast cell disorders | 5 (1) | 3 (1) | 1 |
| ME/CFS (Myalgic encephalomyelitis/chronic fatigue syndrome) | 18 (4) | 9 (4) | 1 |
| Migraines | 90 (20) | 46 (19) | 1 |
| Neurologic conditions (including seizures, dementia, multiple sclerosis, Parkinson’s, neuropathy, small fiber neuropathy, etc.) | 21 (5) | 9 (4) | 1 |
| Postural orthostatic tachycardia syndrome (POTS) or dysautonomia | 14 (3) | 8 (3) | 1 |
| Spinal disorder(s) | 17 (4) | 9 (4) | 1 |
| Tremors/Internal vibrations | 6 (1) | 6 (3) | 1 |
| Depressive disorders | 128 (29) | 49 (20) | 0.66 |
| Anxiety disorders | 135 (31) | 61 (25) | 1 |
| Schizophrenia spectrum and other psychotic disorders | 1 (0) | 0 (0) | 1 |
| Bipolar and related disorders | 15 (3) | 1 (0) | 0.56 |
| Obsessive-Compulsive and related disorders | 15 (3) | 6 (3) | 1 |
| Trauma- and stressor-related disorders | 47 (11) | 17 (7) | 1 |
| Feeding and eating disorders | 21 (5) | 5 (2) | 1 |
| Somatic symptoms (excessive thoughts, feelings, and behaviors relating to the physical symptoms) and related disorders | 7 (2) | 2 (1) | 1 |

^1^n/N, % for all variables

^2^Wilcoxon rank sum test; Fisher's exact test; Pearson's Chi-squared test

LC, long COVID; PVS, Post-vaccination syndrome

**eTable 2. Participants’ health status.**

| **Characteristic** | **LC**  **N = 441** | **PVS**  **N = 241** | **P-value*^4^*** |
| --- | --- | --- | --- |
| Euro-QoL visual analogue scale (0-100)^1, 2^ | 49 (32-61) | 50 (39-70) | 0.02 |
| How bad are your LC/PVS or other symptoms (0-100) on your worst days?*^1, 3^* | 79 (65-86) | 80 (69-89) | 0.07 |
| Missing | 26 | 3 |  |
| Self-reported health status*^1^* |  |  | 0.22 |
| Don’t know | 16 (4) | 15 (6) |  |
| Excellent | 29 (7) | 17 (7) |  |
| Fair | 129 (30) | 48 (20) |  |
| Good | 81 (19) | 56 (23) |  |
| Poor | 108 (25) | 58 (24) |  |
| Very good | 78 (18) | 47 (20) |  |

^1^Median (IQR) EQ-VAS and symptom severity; n/N, % for self-reported health status

^2^The Euro-QoL visual analogue scale is measured from 0 (worst imaginable health) to 100 (best imaginable health).

^3^Symptom severity is measured from 0 (trivial illness) to 100 (unbearable).

^4^Wilcoxon rank sum test for EQ-VAS and symptom severity; Pearson's Chi-squared test for self-reported health status

LC, long COVID; PVS, post-vaccination syndrome

**eTable 3. New-onset conditions.**

| **New-onset Condition** | **LC (%)**  **N = 441*^1^*** | **PVS (%)**  **N = 241*^1^*** | **P-value*^2^*** |
| --- | --- | --- | --- |
| Any allergies | 192 (44) | 108 (45) | 1 |
| Arthritis (including rheumatoid arthritis, gout, lupus, or fibromyalgia) | 98 (22) | 42 (17) | 1 |
| Asthma | 84 (19) | 37 (15) | 1 |
| Autoimmune disease (including lupus, scleroderma, etc.) | 71 (16) | 47 (20) | 1 |
| Bleeding disorder (including sickle cell disease or Thalassemia) | 6 (1) | 2 (1) | 1 |
| Blood clots | 17 (4) | 10 (4) | 1 |
| Cancer or malignancy of any kind | 14 (3) | 7 (3) | 1 |
| Cerebrovascular conditions affecting blood vessels to or in the brain (including stroke) | 9 (2) | 13 (5) | 0.88 |
| Chronic lung disease (including emphysema, chronic bronchitis, chronic obstructive pulmonary disease (COPD), or pulmonary fibrosis) | 24 (5) | 5 (2) | 1 |
| Cystic fibrosis | 0 (0) | 0 (0) | 1 |
| Diabetes | 17 (4) | 6 (3) | 1 |
| Ehlers-Danlos Syndrome (hypermobile joints) | 18 (4) | 15 (6) | 1 |
| Gastrointestinal issues (including IBS or acid reflux) | 144 (33) | 73 (30) | 1 |
| Heart attack, also called myocardial infarction | 2 (0) | 3 (1) | 1 |
| Heart conditions (including coronary artery disease or cardiomyopathies) | 43 (10) | 31 (13) | 1 |
| Heart failure | 1 (0) | 3 (1) | 1 |
| High cholesterol | 83 (19) | 52 (22) | 1 |
| History of organ transplant (including kidney, liver, heart, or lung) | 0 (0) | 0 (0) | 1 |
| Hypertension or high blood pressure | 75 (17) | 45 (19) | 1 |
| Immunocompromised state (including weakened immune system from blood or bone marrow transplant, immune deficiencies, HIV, use of corticosteroids, or use of other immune-weakening medicines) | 30 (7) | 22 (9) | 1 |
| Kidney disease | 8 (2) | 2 (1) | 1 |
| Liver disease | 4 (1) | 6 (3) | 1 |
| Lyme disease | 11 (2) | 9 (4) | 1 |
| MCAS (mast cell activation syndrome) or other mast cell disorders | 29 (7) | 28 (12) | 1 |
| ME/CFS (Myalgic encephalomyelitis/chronic fatigue syndrome) | 82 (19) | 28 (12) | 0.85 |
| Migraines | 109 (25) | 53 (22) | 1 |
| Neurologic conditions (including seizures, dementia, multiple sclerosis, Parkinson’s, neuropathy, small fiber neuropathy, etc.) | 72 (16) | 79 (33) | < 0.0001 |
| Postural orthostatic tachycardia syndrome (POTS) or dysautonomia | 122 (28) | 70 (29) | 1 |
| Spinal disorder(s) | 17 (4) | 13 (5) | 1 |
| Depressive disorders | 125 (28) | 49 (20) | 1 |
| Anxiety disorders | 164 (37) | 86 (36) | 1 |
| Schizophrenia spectrum and other psychotic disorders | 2 (0) | 0 (0) | 1 |
| Bipolar and related disorders | 10 (2) | 1 (0) | 1 |
| Obsessive-Compulsive and related disorders | 12 (3) | 6 (3) | 1 |
| Trauma- and stressor-related disorders | 63 (14) | 25 (10) | 1 |
| Feeding and eating disorders | 10 (2) | 4 (2) | 1 |
| Somatic symptoms (excessive thoughts, feelings, and behaviors relating to the physical symptoms) and related disorders | 14 (3) | 4 (2) | 1 |

^1^n/N, % for all variables

^2^Wilcoxon rank sum test; Fisher's exact test; Pearson's Chi-squared test

LC, long COVID; PVS, post-vaccination syndrome

**eTable 4. New treatments.**

| **Treatment** | **LC (%)**  **N = 441*^1^*** | **PVS (%)**  **N = 241*^1^*** | **p-value*^2^*** |
| --- | --- | --- | --- |
| Acetaminophen [Paracetamol] (Tylenol, Panadol, Apra, FeverAll, Mapap, Pharbetol) | 225 (51) | 111 (46) | 1 |
| Acetazolamide (Diamox) | 0 (0) | 3 (1) | 1 |
| Acetyl-L-Carnitine | 47 (11) | 30 (12) | 1 |
| Acyclovir (Zovirax) | 10 (2) | 10 (4) | 1 |
| Alpha lipoic acid | 63 (14) | 62 (26) | 0.06 |
| Alprazolam (Xanax) | 39 (9) | 15 (6) | 1 |
| Amiodarone (Nexterone) | 1 (0) | 0 (0) | 1 |
| Amitriptyline (Elavil) | 28 (6) | 15 (6) | 1 |
| Amlodipine (Norvasc) | 12 (3) | 5 (2) | 1 |
| Antidiarrheal (Dyphenoxilate [Lomotil], Loperamide [Immodium], Bismuth subsalicylate [Kaopectate, Pepto Bismol]) | 56 (13) | 10 (4) | 0.044 |
| Anti-nausea medication (Ondansetron [Zofran], Prochlorperazine [Compazine], Promethazine [Phenergan], Metoclopramide [Reglan]) | 69 (16) | 25 (10) | 1 |
| Antispasmodics (Hyoscyamine [Levsin], Dicyclomine [Bentyl], Clidinium and chlordiazepoxide [Librax], atropine, scopolamine, and phenobarbital [Donnatol]) | 20 (5) | 7 (3) | 1 |
| Direct oral anticoagulant: Apixaban (Eliquis) | 12 (3) | 2 (1) | 1 |
| Aspirin [Acetylsalicylic acid] (Ascriptin, Aspercin, Aspirtab, Buffasal, Bufferin, Buffinol, Easprin, Ecotrin, Vazalore), 81 mg (baby aspirin), 325 mg (regular aspirin) | 144 (33) | 84 (35) | 1 |
| Astaxanthin | 11 (2) | 7 (3) | 1 |
| Atorvastatin (Lipitor) | 29 (7) | 17 (7) | 1 |
| Ayahuasca | 1 (0) | 0 (0) | 1 |
| Ayurveda medicine | 7 (2) | 7 (3) | 1 |
| Baclofen (Lioresal) | 12 (3) | 8 (3) | 1 |
| Bamlanivimab and etesevimab | 1 (0) | 2 (1) | 1 |
| BC 007 | 0 (0) | 0 (0) | 1 |
| Bebtelovimab | 5 (1) | 0 (0) | 1 |
| Direct oral anticoagulant: Betrixaban (Bevyxxa) | 0 (0) | 0 (0) | 1 |
| Black seed oil | 30 (7) | 43 (18) | 0.003 |
| Botulinum toxin injection (Botox) | 16 (4) | 9 (4) | 1 |
| Bronchodilators (Albuterol, Salmeterol, Formoterol, Ipratropium, Tiotropium, other) | 143 (32) | 26 (11) | < 0.0001 |
| Bumetanide (Bumex) | 0 (0) | 0 (0) | 1 |
| Candesartan (Atacand) | 2 (0) | 4 (2) | 1 |
| Cannabis | 85 (19) | 33 (14) | 1 |
| Capsaicin | 14 (3) | 8 (3) | 1 |
| Carbamazepine (Tegretol) | 1 (0) | 2 (1) | 1 |
| CBD (oral, topical, inhaled) | 137 (31) | 68 (28) | 1 |
| Celecoxib (Celebrex, Elyxyb) | 12 (3) | 8 (3) | 1 |
| Cephalosporins (Cephalexin, Ceftriaxone, other) | 11 (2) | 2 (1) | 1 |
| Cetirizine (Zyrtec) | 147 (33) | 87 (36) | 1 |
| Chinese medicine treatments: Acupuncture | 95 (22) | 60 (25) | 1 |
| Chinese medicine treatments: Herbs | 47 (11) | 25 (10) | 1 |
| Chiropractic treatment | 68 (15) | 44 (18) | 1 |
| Chloroquine (Aralen) | 0 (0) | 0 (0) | 1 |
| Chlorpheniramine (Chlor-trimeton) | 8 (2) | 3 (1) | 1 |
| Chlortalidone (Thalitone) | 0 (0) | 0 (0) | 1 |
| Cimetidine (Tagamet) | 10 (2) | 4 (2) | 1 |
| Citalopram (Celexa) | 10 (2) | 5 (2) | 1 |
| Clonazepam (Klonopin) | 22 (5) | 19 (8) | 1 |
| Clopidogrel (Plavix) | 17 (4) | 7 (3) | 1 |
| Cognitive behavioral therapy (e.g. for tinnitus, hearing disorders) | 23 (5) | 13 (5) | 1 |
| Colchicine (Colcrys, Mitigare, Gloperba) | 17 (4) | 14 (6) | 1 |
| Convalescent plasma | 0 (0) | 0 (0) | 1 |
| Copper | 24 (5) | 12 (5) | 1 |
| CoQ10 | 142 (32) | 74 (31) | 1 |
| Cranio-sacral massage | 36 (8) | 24 (10) | 1 |
| Cromoglicic acid (Cromolyn) oral, nasal spray | 29 (7) | 20 (8) | 1 |
| Cyclobenzaprine (Flexeril) | 33 (7) | 22 (9) | 1 |
| Direct oral anticoagulant: Dabigatran (Pradaxa) | 0 (0) | 1 (0) | 1 |
| Dandelion | 18 (4) | 32 (13) | 0.005 |
| DAO enzyme supplement (OTC) | 16 (4) | 19 (8) | 1 |
| Desvenlafaxine (Pristiq) | 7 (2) | 1 (0) | 1 |
| Diazepam (Valium) | 19 (4) | 6 (2) | 1 |
| Lifestyle changes: Diet change | 153 (35) | 95 (39) | 1 |
| Diets: Intermittent fasting | 108 (24) | 95 (39) | 0.016 |
| Diets: Low histamine diet | 100 (23) | 83 (34) | 0.23 |
| Diets: Low salt diet | 29 (7) | 18 (7) | 1 |
| Diets: Other (please describe) | 78 (18) | 55 (23) | 1 |
| Diltiazem (Cardizem) | 9 (2) | 5 (2) | 1 |
| Dimenhydrinate (Dramamine) | 17 (4) | 11 (5) | 1 |
| Diphenhydramine (Benadryl) | 104 (24) | 67 (28) | 1 |
| Doxepin (Sinequan) | 1 (0) | 2 (1) | 1 |
| Doxycycline | 31 (7) | 17 (7) | 1 |
| Duloxetine (Cymbalta) | 37 (8) | 22 (9) | 1 |
| Eastern white pine needle | 3 (1) | 19 (8) | < 0.001 |
| Eculizumab (Soliris) | 0 (0) | 0 (0) | 1 |
| Direct oral anticoagulant: Edoxaban (Savaysa) | 0 (0) | 0 (0) | 1 |
| Enhanced External Counterpulsation (EECP) | 3 (1) | 1 (0) | 1 |
| Enalapril (Epaned) | 1 (0) | 0 (0) | 1 |
| Escitalopram (Lexapro) | 46 (10) | 18 (7) | 1 |
| Tixagevimab and cilgavimab (Evusheld) | 1 (0) | 4 (2) | 1 |
| Famciclovir (Famvir) | 7 (2) | 3 (1) | 1 |
| Famotidine (Pepcid) | 166 (38) | 91 (38) | 1 |
| Fexofenadine (Allegra) | 60 (14) | 39 (16) | 1 |
| Flavenoids (Quercetin, Luteolin, Braingain) | 85 (19) | 71 (29) | 0.64 |
| Flecainide (Tambocor) | 1 (0) | 2 (1) | 1 |
| Fluoroquinolones (Ciprofloxacin, Levofloxacin, other) | 16 (4) | 2 (1) | 1 |
| Fluoxetine (Prozac) | 19 (4) | 8 (3) | 1 |
| Fluvoxamine (Luvox) | 20 (5) | 16 (7) | 1 |
| Furosemide (Lasix) | 6 (1) | 2 (1) | 1 |
| Gabapentin (Neurontin, Gralise) | 87 (20) | 61 (25) | 1 |
| Glutamine | 16 (4) | 10 (4) | 1 |
| Glutathione | 54 (12) | 47 (20) | 1 |
| Hyperbaric Oxygen Therapy (HBOT) | 26 (6) | 15 (6) | 1 |
| Hearing devices/maskers for tinnitus | 12 (3) | 11 (5) | 1 |
| Heparin | 6 (1) | 3 (1) | 1 |
| Home oxygen | 13 (3) | 6 (2) | 1 |
| Lifestyle changes: Hydration, increase or decrease salt intake | 200 (45) | 105 (44) | 1 |
| Hydrochlorothiazide (Microzide) | 8 (2) | 3 (1) | 1 |
| Hydroxychloroquine (Plaquenil) | 27 (6) | 19 (8) | 1 |
| Hydroxyzine (Atarax) (Rx) | 38 (9) | 22 (9) | 1 |
| Ibuprofen (Advil, Motrin, Provil) | 253 (57) | 129 (54) | 1 |
| IL-1 antagonist (Anakinra [Kineret], Canakinumab [Ilaris], other) | 0 (0) | 2 (1) | 1 |
| IL-6 antagonist (Siltuximab [Sylvant], Sarilumab [Kevzara], Tocilizumab [Actemra], other) | 0 (0) | 0 (0) | 1 |
| Inhaled steroids (Mometasone [Asmanex], Fluticasone [Flovent], Budesonide [Pulmicort], other) | 123 (28) | 24 (10) | < 0.0001 |
| Ivermectin (Stromectol) | 44 (10) | 44 (18) | 0.57 |
| Intravenous Immunoglobulin (IVIG) | 7 (2) | 17 (7) | 0.08 |
| Integrative medicine treatments: IV ozone | 12 (3) | 7 (3) | 1 |
| IV steroids (Hydrocortisone, Methylprednisolone, other) | 12 (3) | 12 (5) | 1 |
| Integrative medicine treatments: IV vitamins | 34 (8) | 25 (10) | 1 |
| Ketamine (Ketalar) | 5 (1) | 2 (1) | 1 |
| Ketorolac (Toradol) | 10 (2) | 6 (2) | 1 |
| Ketotifen (Zaditor) | 8 (2) | 12 (5) | 1 |
| Kinase inhibitors (Acalabrutinib [Calquence], Ibrutinib [Imbruvica], Zanubrutinib [Brukinsa], Baricitinib [Olumiant], Ruxolitinib [Jakafi], Tofacitinib [Xeljanz], other) | 2 (0) | 1 (0) | 1 |
| Lamotrigine (Lamictal) | 8 (2) | 2 (1) | 1 |
| Laxatives, stool softeners | 65 (15) | 32 (13) | 1 |
| Lidocaine | 41 (9) | 16 (7) | 1 |
| Lifestyle changes: Limiting exercise or exertion | 284 (64) | 124 (51) | 0.23 |
| Lisinopril (Prinivil, Zestril) | 15 (3) | 7 (3) | 1 |
| L-Lysine | 42 (10) | 25 (10) | 1 |
| Loratadine (Claritin) | 108 (24) | 84 (35) | 0.92 |
| Lorazepam (Ativan) | 26 (6) | 21 (9) | 1 |
| Losartan (Cozaar) | 17 (4) | 4 (2) | 1 |
| Low dose Naltrexone | 95 (22) | 48 (20) | 1 |
| Lysergic Acid Diethylamide (LSD) | 1 (0) | 0 (0) | 1 |
| Lymphatic massage | 48 (11) | 37 (15) | 1 |
| Macrolides (Azithromycin, Clarithromycin, Erythromycin) | 34 (8) | 7 (3) | 1 |
| Magnesium | 206 (47) | 135 (56) | 1 |
| CCR5 entry inhibitors: Maraviroc (Selzentry) | 17 (4) | 11 (5) | 1 |
| 3, 4-Methylenedioxymethamphetamine (MDMA) | 2 (0) | 0 (0) | 1 |
| Meclizine (Bonine) | 24 (5) | 17 (7) | 1 |
| Melatonin | 187 (42) | 79 (33) | 1 |
| Metaxalone (Skelaxin) | 2 (0) | 1 (0) | 1 |
| Methocarbomal (Robaxin) | 6 (1) | 5 (2) | 1 |
| Metoprolol (Lopressor) | 45 (10) | 24 (10) | 1 |
| Midodrine (Proamatine) | 15 (3) | 7 (3) | 1 |
| Milk thistle | 22 (5) | 27 (11) | 0.99 |
| Mirtazapine (Remeron) | 12 (3) | 2 (1) | 1 |
| Molnupiravir (Lagevrio) | 4 (1) | 0 (0) | 1 |
| Monoclonal immunosuppressives: CD-20/B-Cell modulator | 0 (0) | 1 (0) | 1 |
| Monoclonal immunosuppressives: Rituximab (Riabni, Rituxan, Ruxience, Truxima) | 1 (0) | 0 (0) | 1 |
| Monoclonal immunosuppressives: Leronlimab | 0 (0) | 0 (0) | 1 |
| Montelukast (Singulair) | 50 (11) | 22 (9) | 1 |
| Moringa | 6 (1) | 9 (4) | 1 |
| NAC (N-Acetyl Cysteine)/Glycine | 107 (24) | 79 (33) | 1 |
| Naproxen (Aleve, Mediproxen, All Day Pain Relief) | 110 (25) | 58 (24) | 1 |
| Nasal spray (Flonase [Fluticasone], Nasocort [Triamcinolone], Afrin [Oxymetazoline], other) | 149 (34) | 59 (24) | 1 |
| Nattokinase | 64 (15) | 32 (13) | 1 |
| Nebivolol (Bystolic) | 3 (1) | 2 (1) | 1 |
| Nettles | 13 (3) | 13 (5) | 1 |
| Lifestyle changes: No alcohol or caffeine | 183 (41) | 105 (44) | 1 |
| Nortriptyline (Pamelor) | 10 (2) | 7 (3) | 1 |
| Omalizumab (Xolair) | 6 (1) | 1 (0) | 1 |
| Omega 3s/Fish oil | 161 (37) | 97 (40) | 1 |
| Opioid analgesics (Morphine, Buprenorphine, Codeine, Fentanyl, Oxycodone, Hydrocodone, Hydromorphone, other) | 19 (4) | 10 (4) | 1 |
| Oral (Prednisone, Dexamethasone, Hydrocortisone, Methylprednisolone, other) | 133 (30) | 116 (48) | < 0.001 |
| Other steroid injections (intratympanic, spinal) | 10 (2) | 9 (4) | 1 |
| Paroxetine (Brisdelle, Paxil, Pexeva) | 3 (1) | 1 (0) | 1 |
| Nirmatrelvir/Ritonavir (Paxlovid) | 47 (11) | 4 (2) | 0.001 |
| PEA (Palmitoylethanolamide) | 13 (3) | 17 (7) | 1 |
| Penicillin (Penicillin, Amoxicillin, other) | 25 (6) | 10 (4) | 1 |
| Plasmapheresis/Apheresis | 1 (0) | 2 (1) | 1 |
| Pravastatin (Pravachol) | 20 (5) | 11 (5) | 1 |
| Pregabalin (Lyrica) | 25 (6) | 13 (5) | 1 |
| Probiotics | 419 (95) | 238 (99) | 1 |
| Propranolol (Inderal) | 65 (15) | 27 (11) | 1 |
| Proton pump inhibitors (Omeprazole [Prilosec], Esomeprazole [Nexium]) | 98 (22) | 43 (18) | 1 |
| Psilocin (4-HO-DMT) | 5 (1) | 3 (1) | 1 |
| Physical therapy: Rehabilitation | 94 (21) | 35 (15) | 1 |
| Physical therapy: Speech | 40 (9) | 4 (2) | 0.013 |
| Physical therapy: Strengthening, stamina | 101 (23) | 36 (15) | 1 |
| Physical therapy: Vestibular | 31 (7) | 15 (6) | 1 |
| Physical therapy: Vestibular-ocular | 13 (3) | 6 (2) | 1 |
| Red light therapy | 20 (5) | 27 (11) | 0.3 |
| Casirivimab/Imdevimab (Regeneron) | 5 (1) | 7 (3) | 1 |
| Reishi, Lion’s Mane, Cordyceps (mushrooms) | 52 (12) | 28 (12) | 1 |
| Remdesivir (Veklury) | 3 (1) | 0 (0) | 1 |
| Treatment for SIBO (Rifaximin [Xifaxin], other) | 7 (2) | 4 (2) | 1 |
| Direct oral anticoagulant: Rivaroxaban (Xarelto) | 0 (0) | 4 (2) | 1 |
| Rosuvastatin (Crestor) | 10 (2) | 7 (3) | 1 |
| SaltStick | 31 (7) | 11 (5) | 1 |
| Subcutaneous Immunoglobulin (SCIG) | 0 (0) | 2 (1) | 1 |
| Scopolamine | 3 (1) | 2 (1) | 1 |
| Sertraline (Zoloft) | 40 (9) | 11 (5) | 1 |
| Simethicone (Gas-X) | 51 (12) | 16 (7) | 1 |
| Sotrovimab (Xevudy) | 3 (1) | 0 (0) | 1 |
| Spironolactone (Aldactone, CaroSpir) | 8 (2) | 7 (3) | 1 |
| St. John’s Wort (hypericum perforatum) | 9 (2) | 4 (2) | 1 |
| T-Cell and B-Cell inhibitors (mycophenolate mofetil [CellCept]) | 0 (0) | 2 (1) | 1 |
| Temazepam (Restoril) | 3 (1) | 1 (0) | 1 |
| Tizanidine (Zanaflex) | 15 (3) | 13 (5) | 1 |
| Tramadol (ConZip, Qdolo, Ultram) | 17 (4) | 4 (2) | 1 |
| Trazodone (Desyrel) | 34 (8) | 12 (5) | 1 |
| Triazolam (Halcion) | 0 (0) | 0 (0) | 1 |
| Tumeric (Curcumin) | 162 (37) | 96 (40) | 1 |
| Integrative medicine treatments: Treatment for EBV, CMV, herpes, Lyme disease | 21 (5) | 9 (4) | 1 |
| Valcyclovir (Valtrex) | 55 (12) | 23 (10) | 1 |
| Venlafaxine (Effexor) | 17 (4) | 8 (3) | 1 |
| Verapamil (Calan, Isoptin) | 2 (0) | 4 (2) | 1 |
| Warfarin (Coumadin) | 1 (0) | 0 (0) | 1 |
| Zaleplon (Sonata) | 1 (0) | 0 (0) | 1 |
| Zinc | 154 (35) | 86 (36) | 1 |
| Zolpidem (Ambien) | 27 (6) | 13 (5) | 1 |
| Vision therapy | 14 (3) | 6 (2) | 1 |
| Vitamin B1 (Thiamine) | 70 (16) | 52 (22) | 1 |
| Vitamin B12 (hydroxocobalamin, cyanocobalamin) | 160 (36) | 103 (43) | 1 |
| Vitamin B2 (Riboflavin) | 66 (15) | 42 (17) | 1 |
| Vitamin B3 (Niacin) | 79 (18) | 36 (15) | 1 |
| Vitamin B6 (Pyridoxine) | 48 (11) | 39 (16) | 1 |
| Vitamin B complex | 150 (34) | 102 (42) | 1 |
| Vitamin C (ascorbic acid) | 184 (42) | 128 (53) | 1 |
| Vitamin D (calciferol) | 287 (65) | 157 (65) | 1 |
| Vitamin E (alpha-tocopherol) | 50 (11) | 22 (9) | 1 |
| Vitamin K2 (menaquinone) | 50 (11) | 52 (22) | 0.1 |

^1^n/N, % for all variables

^2^Wilcoxon rank sum test; Fisher's exact test; Pearson's Chi-squared test

LC, long COVID; PVS, post-vaccination syndrome

**eFigure 1. Study flow diagram.**

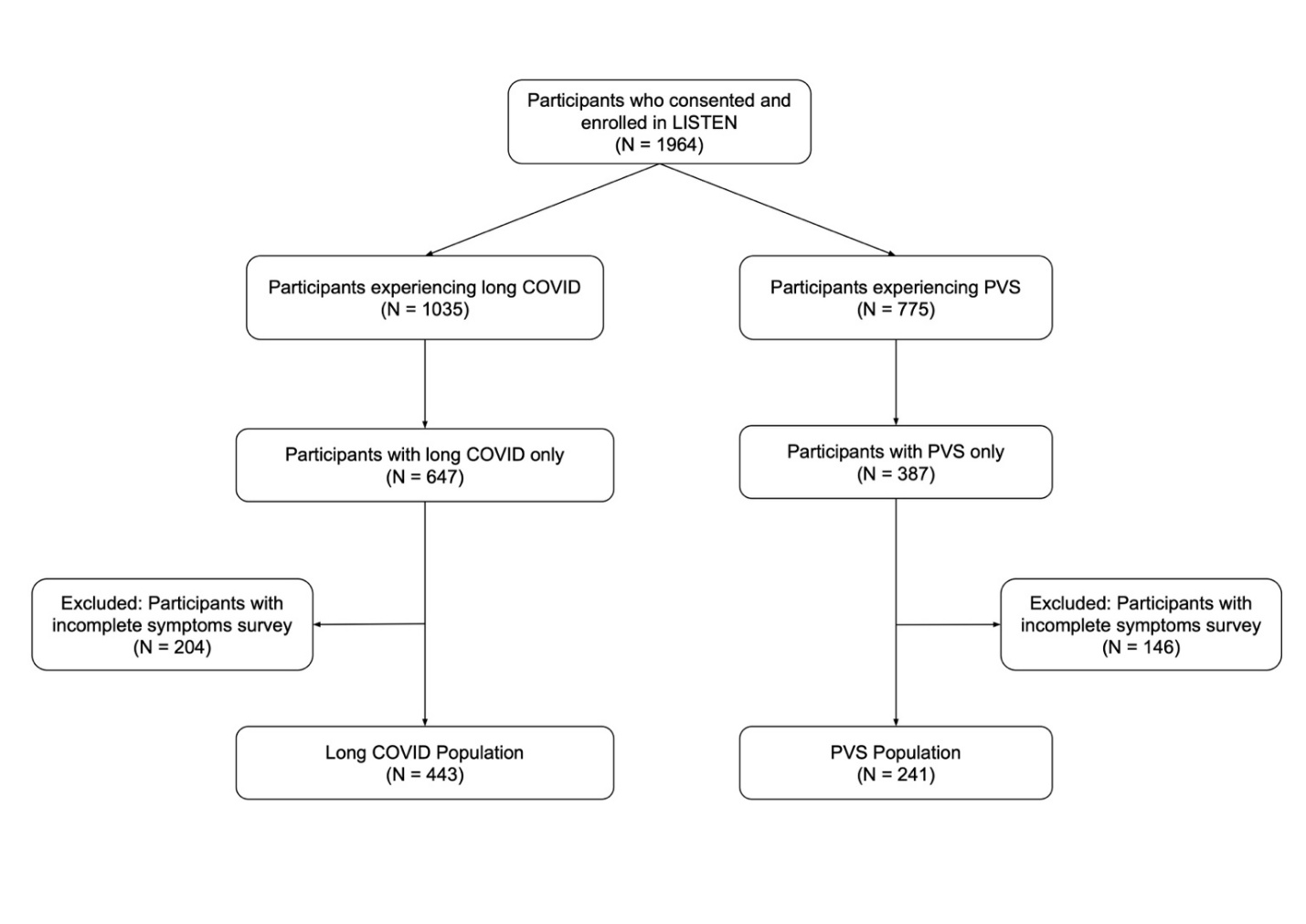

LISTEN, Listen to Immune, Symptom and Treatment Experiences Now; PVS, post-vaccination syndrome

**eFigure 2. Distribution of health status (A) measured by the Euro-QoL visual analogue scale and stratified by reported condition and (B) measured by symptom severity and stratified by reported condition.**

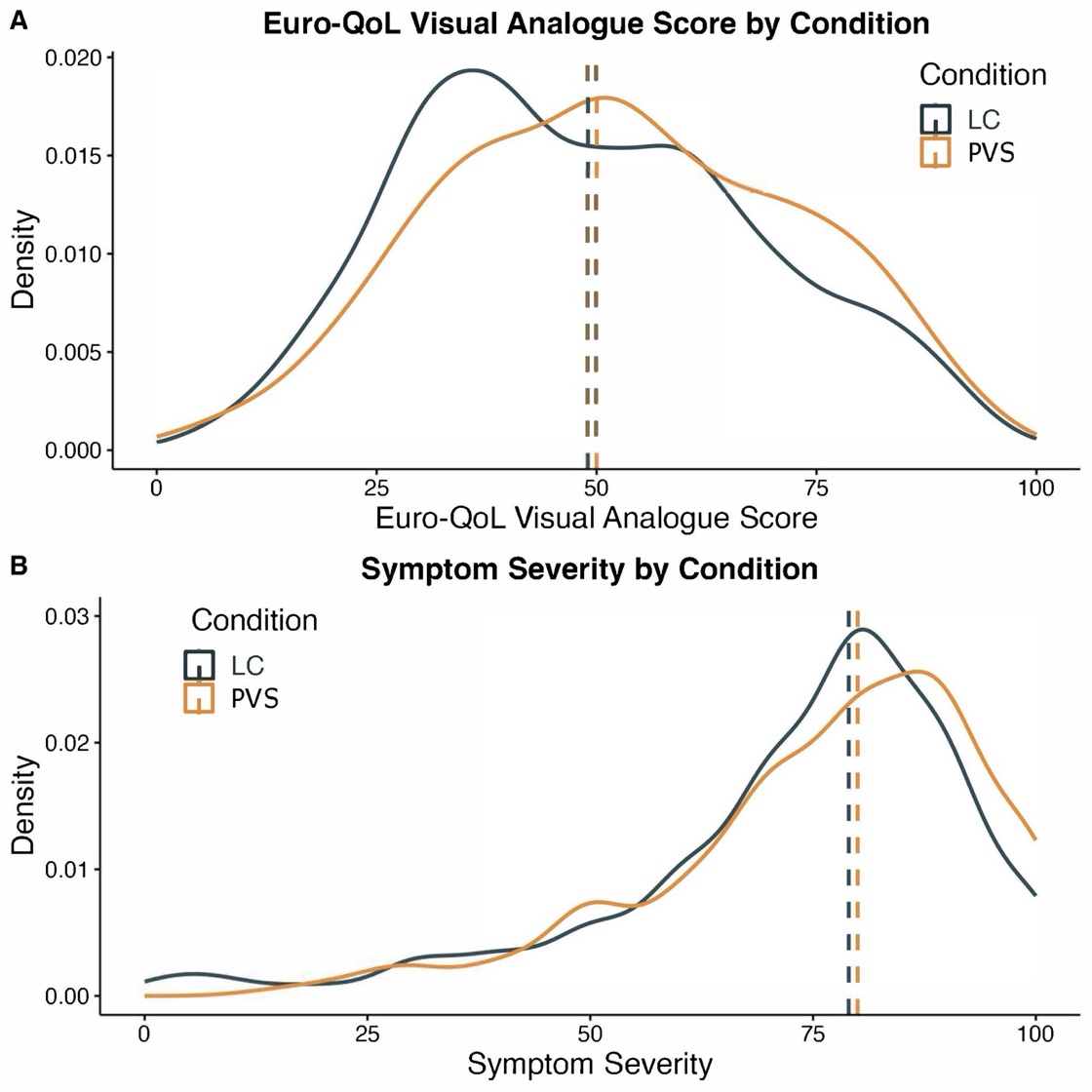

Median values are indicated with dashed lines. EQ-VAS was assessed by the question, “Please choose one point in this 0-100 scale, which can best represent your health today (0 means the worst and 100 means the best).” Symptom severity was assessed by the question, “We are trying to get a sense of how bad your PVS/LC symptoms are when you feel them the most. On the slider below, with 0 being a trivial illness and 100 being unbearable, please let us know what the worst days are like.”

LC, long COVID; PVS, post-vaccination syndrome

**eFigure 3. Differences in reported symptoms between long COVID and PVS.**

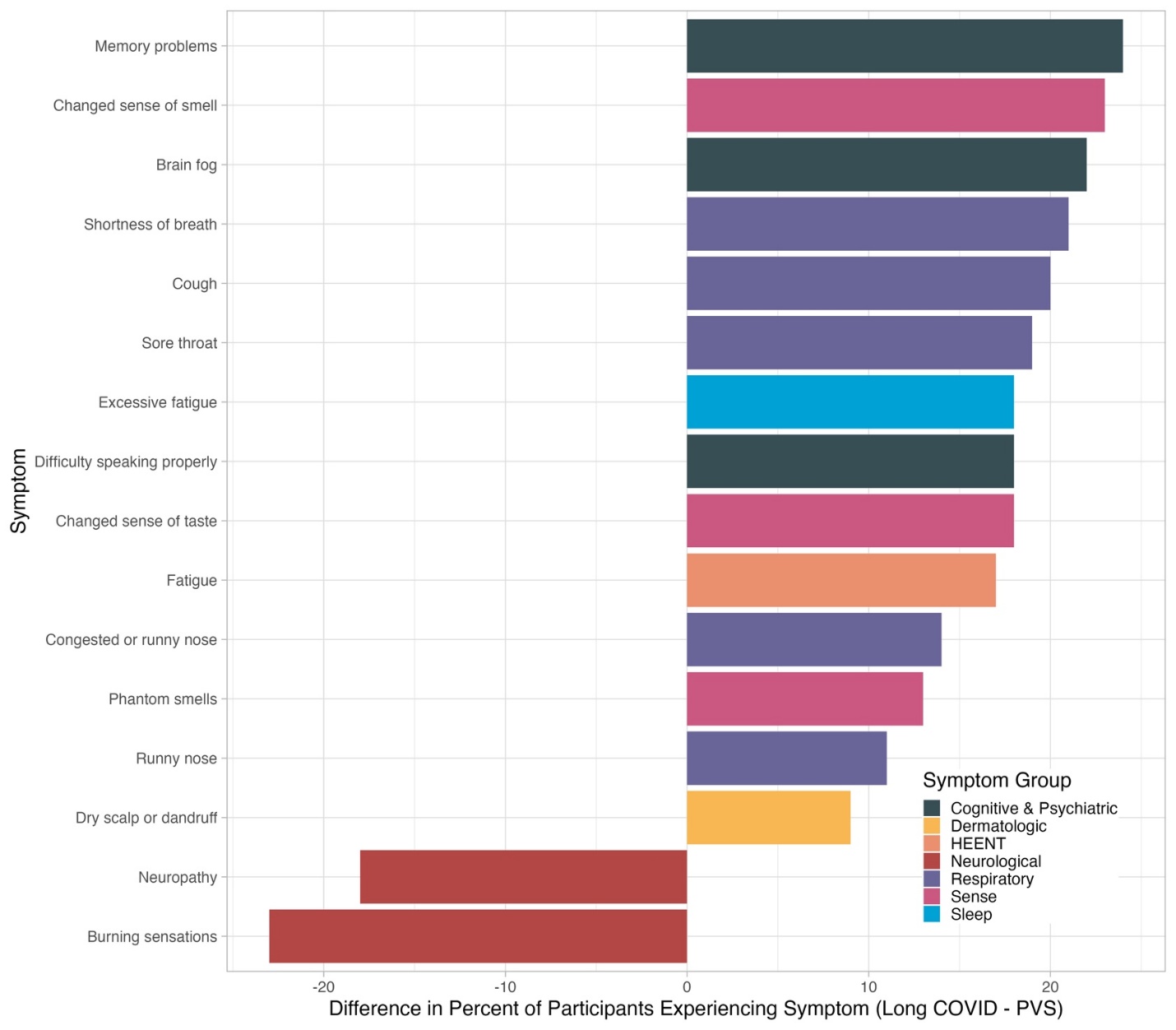

HEENT, head, eyes, ears, nose, and throat; PVS, post-vaccination syndrome

**eFigure 4. Differences in treatment use between long COVID and PVS.**

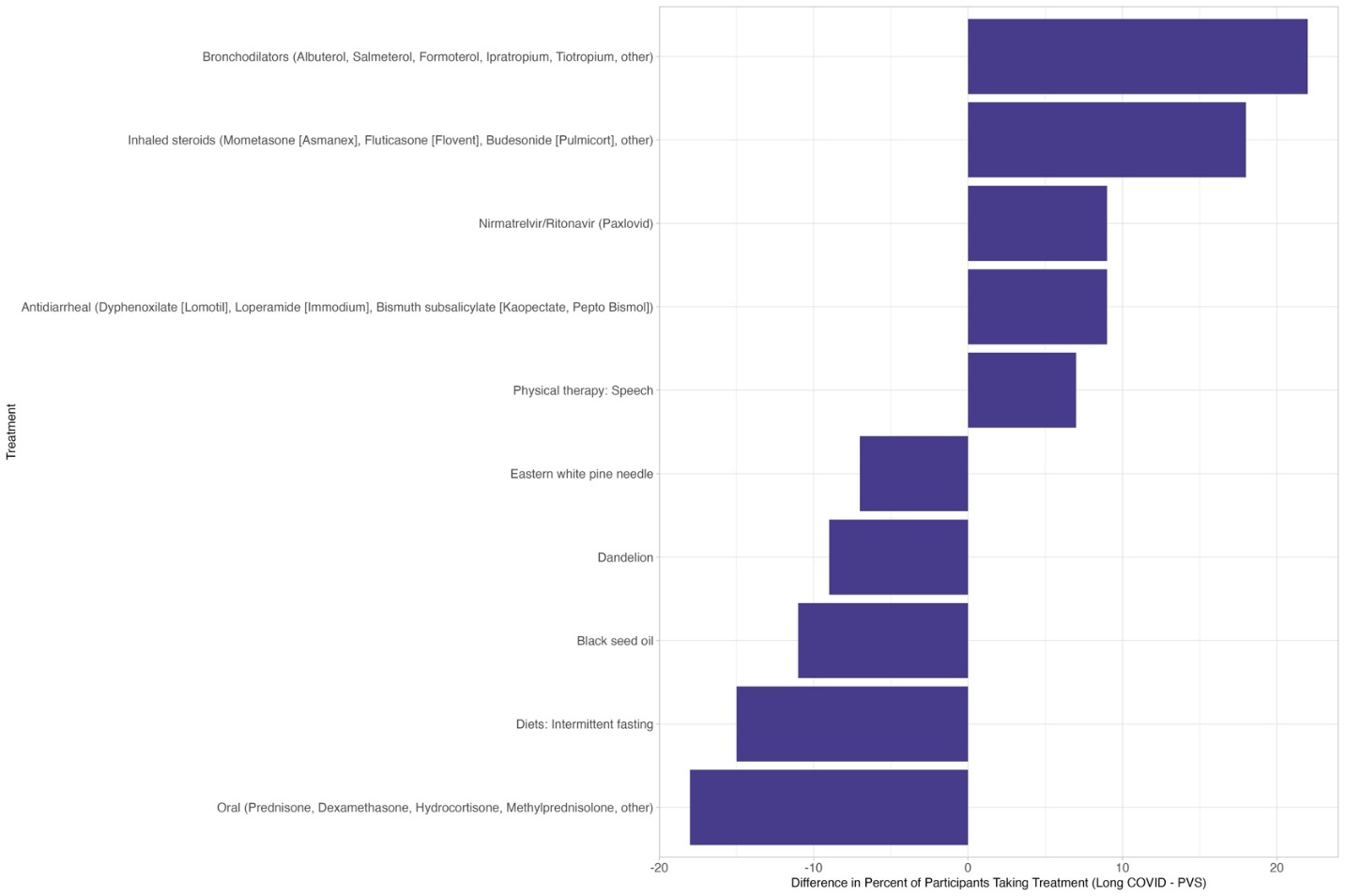

PVS, post-vaccination syndrome
